# Lipoprotein (a) is associated with increased risk of Abdominal Aortic Aneurysm

**DOI:** 10.1101/2024.09.13.24313646

**Authors:** Pranav Sharma, Renae Judy, Shuai Yuan, Corry Gellatly, Katie L. Saxby, Matthew J. Bown, Michael G. Levin, Scott M. Damrauer

## Abstract

**Introduction:** Lipoprotein(a) (Lp(a)) is a circulating apolipoprotein B (ApoB) containing particle that has been observationally linked to atherosclerotic cardiovascular disease and is the target of emerging therapeutics. Recent work has highlighted the role of circulating lipoproteins in abdominal aortic aneurysm (AAA). We sought to triangulate human observational and genetic evidence to evaluate the role of Lp(a) in AAA.

**Methods:** We tested the association between circulating levels of Lp(a) and clinically diagnosed abdominal aortic aneurysms while controlling for traditional AAA risk factors and levels of ApoB using logistic regression among 795 individuals with and 374,772 individuals without AAA in the UK Biobank (UKB). Multivariable Mendelian randomization (MVMR) was used to test for putatively causal associations between Lp(a) and AAA controlling for ApoB. Genetic instruments for Lp(a) and ApoB were created from genome-wide association studies (GWAS) of Lp(a) and ApoB comprising 335,796 and 418,505 UKB participants, respectively. The instruments were tested for association with AAA using data from a GWAS of 39,221 individuals with and 1,086,107 without AAA.

**Results:** Elevated Lp(a) levels were observationally associated with an increased risk of AAA (OR 1.04 per 10 nmol/L Lp(a); 95%CI 1.02-1.05; P<0.01). Clinically elevated Lp(a) levels (>150nmol/L) were likewise associated with an increased risk of AAA (OR 1.47; 95% CI 1.15-1.88; P < 0.01) when compared to individuals with Lp(a) levels <150nmol/L. MVMR confirmed a significant, ApoB-independent association between increased Lp(a) and increased risk of AAA (OR 1.13 per SD increase in Lp(a); 95%CI 1.02-1.24; P<0.02).

**Conclusion:** Both observational and genetic analyses support an association between increased Lp(a) and AAA risk that is independent of ApoB. These findings suggest that Lp(a) may be a therapeutic target for AAA and drive the inclusion of AAA as an outcome in clinical trials of Lp(a) antagonists.

## INTRODUCTION

Abdominal aortic aneurysm (AAA) is a life-threatening condition in which progressive dilatation of the infrarenal aorta leads to rupture. Abdominal aortic aneurysms are have been linked to 1.3% of all deaths among men aged 65-85.^1^ While historical prevalence estimates ranged from 3.9% to 7.9%, more recent figures suggest rates between 1.2% and 3.3% in individuals 60 years or older.^2^

Current guidelines recommend screening via duplex ultrasound in men and women aged 65-75 years old who have a family history of AAA or have ever smoked.^3^ Because AAAs grow slowly over time, and rupture risk is proportional to aneurysm size, the mainstay of management is longitudinal monitoring by aortic duplex ultrasound until aneurysm size reaches the point at which the risk of rupture exceeds the risk of repair.^4^ Multiple pharmacological therapies have been proposed for AAA based on compelling biology and promising evidence from preclinical model systems, including ACE inhibitors, angiotensin receptor blockers, and matrix metalloproteinase inhibitors. None of these have been shown to affect aneurysm growth or rupture in human trials and they are not currently recommended for the treatment of AAA.^5–11^

Recent large scale efforts have identified inflammation and atherogenesis as key processes underlying the development of AAA, and identified genetic variants at the lipoprotein (a) locus that confer increased susceptibility to AAA.^12,13^ Lipoprotein(a) (Lp(a)) is an apolipoprotein B (ApoB) containing molecule that has been observationally associated with atherosclerotic cardiovascular disease through putative mechanisms including inflammation, thrombosis, calcification, and atherogenesis.^14^ Lp(a) is one of the most heritable human traits, and is strongly associated with multiple atherosclerotic cardiovascular diseases (ASCVD).^14^ With the emergence of therapies targeting Lp(a),^15^ we sought to further explore this relationship in the context of AAA pathogenesis.

We sought to investigate the role of Lp(a) in AAA pathogenesis through both observational and genetic analyses. Leveraging large-scale observational data from biobanks, we examined the relationship between Lp(a) levels and AAA risk, while accounting for established risk factors. Additionally, we utilized genetic evidence, employing multivariable Mendelian randomization (MR) to evaluate the association of Lp(a) with AAA in conjunction with ApoB. MR leverages the natural randomization of alleles to mimic the results of randomized clinical trials, contingent upon key assumptions.^16,17^ Using multivariable MR allows for the simultaneous assessment of multiple exposures, providing a more comprehensive understanding of their independent and combined effects on AAA risk. This comprehensive approach not only enhances our understanding of AAA pathophysiology but also highlights the potential of Lp(a) as a therapeutic target, offering new avenues for non-invasive intervention.

## METHODS

The study comprised two components. The first was an observational analysis to test the association of measured Lp(a) with AAA while controlling for ApoB and traditional AAA risk factors using data from the UK Biobank (UKB). The second was a causal inference experiment, using multivariable MR (MVMR) to test for a putatively causal association between Lp(a) and AAA while controlling for ApoB. Our analysis was performed in accordance with the STROBE-MR guidelines.^18^ All participants in the underlying studies gave written consent to the use of their data and those studies were approved by the relevant IRB.

### Observational analysis of Lp(a) and AAA

#### Study design and setting

We conducted a cross-sectional cohort analysis using data obtained from the UK Biobank, which comprises over 500,000 individuals aged 40 to 69 years old, enrolled between 2004 and 2010.^19^ Our analysis was restricted to the subset of individuals from the UK Biobank with measured levels of Lipoprotein(a) (Lp(a)) and Apolipoprotein B (ApoB).

Serum Lp(a) concentration, as a continuous, raw measurement was the primary exposure. Serum Lp(a) concentrations were measured using an immuno-turbidimetric assay (Assay Manufacturer: Randox Bioscience, UK; Analytic Platform: Beckman Coulter AU5800) with an analytic range of 5.76-189 nmol/L.^20^ We also tested the association of clinically elevated levels of Lp(a), above 150nmol/L, as defined by the inclusion criteria of the ongoing Lp(a)HORIZON clinical trial,^21^ with AAA.

Covariates included ApoB, which was measured using an immuno-turbidimetric assay (Assay Manufacturer: Beckman Coulter, UK, Ltd; Analytic Platform: Beckman Coulter AU5800) with an analytic range of 0.4-2 g/L, age, sex, race/ethnicity, diabetes status, smoking status, hypertension, hyperlipidemia, and lipid-lowering medication use. Data fields from the UKB data dictionary can be found in Supplementary Table 1.

#### Primary outcome

The primary outcome was prevalent abdominal aortic aneurysm as indicated by at least one instance of an applicable ICD-9 (International Classification of Disease version 9), ICD-10 or OPCS-4 (OPCS Classification of Interventions and Procedures). ICD-9 codes included 441.3 or 441.4. ICD-10 codes included I71.3 or I71.4. OPCS-4 codes included L184, L185, L186, L194, L195, L196, L271, L275, L276, L281, L285, L286. Data fields from the UKB data dictionary can be found in **Supplementary Table 1.**

#### Statistical Methods

We used generalized linear regression to test the relationship between Lp(a) concentration and prevalent AAA. Initially, we examined a model that assessed AAA risk using Lp(a) adjusted for age, sex, and race/ethnicity. Subsequently, we extended our analysis to include additional covariates such as ApoB concentration, diabetes, smoking, hypertension, hyperlipidemia, and lipid-lowering medications. We report the odds ratios of prevalent AAA for a 10nmol/L increase in Lp(a).

Next, we explored the relationship of individuals with clinically elevated levels of Lp(a), above 150nmol/L, as defined by the inclusion criteria of the ongoing Lp(a)HORIZON clinical trial.^21^ We report the odds ratio of individuals with clinically elevated levels of Lp(a) (>150nmol/L) compared to those with normal levels (<150nmol/L), while adjusting for covariates as above.

Finally, to further investigate the risk associated with varying Lp(a) levels, we categorized Lp(a) into 10 nmol/L bins, using the median Lp(a) concentration as reference. This approach, while controlling for covariates, allowed us to visualize the risk of AAA across different Lp(a) levels, providing insight into whether the relationship was linear or exhibited a threshold effect at higher concentrations. We report the odds ratio of bins at high levels of Lp(a) (>150nmol/L) when compared to the median concentration of Lp(a) in this population.

All statistical analyses were computed using R Statistical Software (RStudio 2023.09.1+494 "Desert Sunflower" Release). Statistical significance was determined based on a P-value < 0.05.

### Genetic Causal Inference

#### Study Design and Setting

Multivariable MR was used to test the association between genetically determined Lp(a) levels and genetic liability to AAA, while controlling for genetically determined ApoB levels. Publicly available summary statistics were obtained from the largest genome-wide association studies of these traits.^12,22^

#### Mendelian Randomization Assumptions

A genetic variant can be considered as an instrumental variable for a given exposure if it satisfies the instrumental variable assumptions: 1) it is associated with the exposure, 2) it is not associated with the outcome due to alternative confounding pathways, and 3) it does not affect the outcome except potentially through the exposure.^23,24^ By using large GWAS and evaluating r^2^ and F-statistic of the instruments, we can ensure that the first assumption is met. The second and third assumptions cannot be affirmatively proven; however, by performing a range of pleiotropy-robust sensitivity analyses, these assumptions may be falsified.

#### Genetic Data

The primary exposure was Lp(a); we also tested the association of ApoB independently and in a joint model. Genetic instruments for Lp(a) and ApoB were developed from previously published GWAS data from the Pan-UKB project, comprising 335,796 and 418,505 UK Biobank participants, respectively.^22^ Summary statistics are publicly available at https://pan.ukbb.broadinstitute.org/downloads.^22^

The primary outcome was prevalent AAA.^12^ Summary statistics for AAA were obtained from the recently published multi-population GWAS meta-analysis by Roychowdhury et al. comprising 39,221 individuals with and 1,086,107 without AAA from 14 cohorts. GWAS summary statistics for AAA are publicly available at https://csg.sph.umich.edu/willer/public/AAAgen2023/.

#### Exposure Instrument Selection

Independent (r^2^<0.001, greater than 10,000 kb) genetic variants associated with either Lp(a) or Apo(B) at genome-wide significance (P<5×10^-8^) were used to instrument the exposures. F-statistics and cumulative r^2^ were calculated for each instrument to evaluate the strength of association between the instrument and exposure.

#### Statistical Methods

We first performed a univariable analysis to test the association between Lp(a) and AAA using inverse-variance weighting with multiplicative random effects. Weighted median, weighted mode, and MR Egger methods were applied as sensitivity analyses; these methods make different assumptions about the presence of pleiotropy, which may invalidate the second and third MR assumptions.^16,17^ Because Lp(a) particles contain ApoB, it is possible that the effects of Lp(a) on AAA could be mediated by ApoB. To account for this possibility, we performed multivariable MR (MVMR) which accounts for pleiotropic effects of Lp(a)-associated variants on ApoB and estimates the direct effect of Lp(a) on AAA. Two-sample Mendelian randomization was performed using the TwoSampleMR package in R.^25–28^ Statistical significance was determined based on a P-value < 0.05; the Bonferroni correction was used to control for multiple testing in the univariate two sample MR experiments.

## RESULTS

### Observational analysis of Lp(a) and AAA

There were 375,567 UKB participants with measured Lp(a) available for analysis. They had a median age of 58 at enrollment (Interquartile range [IQR] 50-63), 171,259 (45.6%) were male, and 194,821 (51.9%) were ever smokers (Table 1). The study population included 795 individuals with AAA and 374,772 individuals without AAA. Using a generalized linear regression model, we first tested for an association between Lp(a) and AAA, controlling for age, sex, and race/ethnicity, finding a 4% increase in risk per 10nmol/L increase in Lp(a) (OR 1.04; 95% CI 1.02-1.05; P < 0.01). This association was robust to controlling for clinical risk factors and covariates, including diabetes, smoking, hypertension, hyperlipidemia, and the use of lipid lowering medications, as well ApoB (OR 1.04 per 10nmol increase of Lp(a); 95% CI 1.02-1.05; P < 0.01). This analysis identified several covariates that exhibited significant associations with AAA risk (P < 0.01), including male sex (OR 13.87; 95% CI 10.02-19.20), hypertension (OR 3.28; 95% CI 2.06-5.22), smoking (OR 3.05; 95% CI 2.43-3.84), and age (OR 1.19; 95% CI 1.17-1.21). Results of all covariates and their association with AAA risk are shown in **Supplementary Table 2**.

**Table 1.**
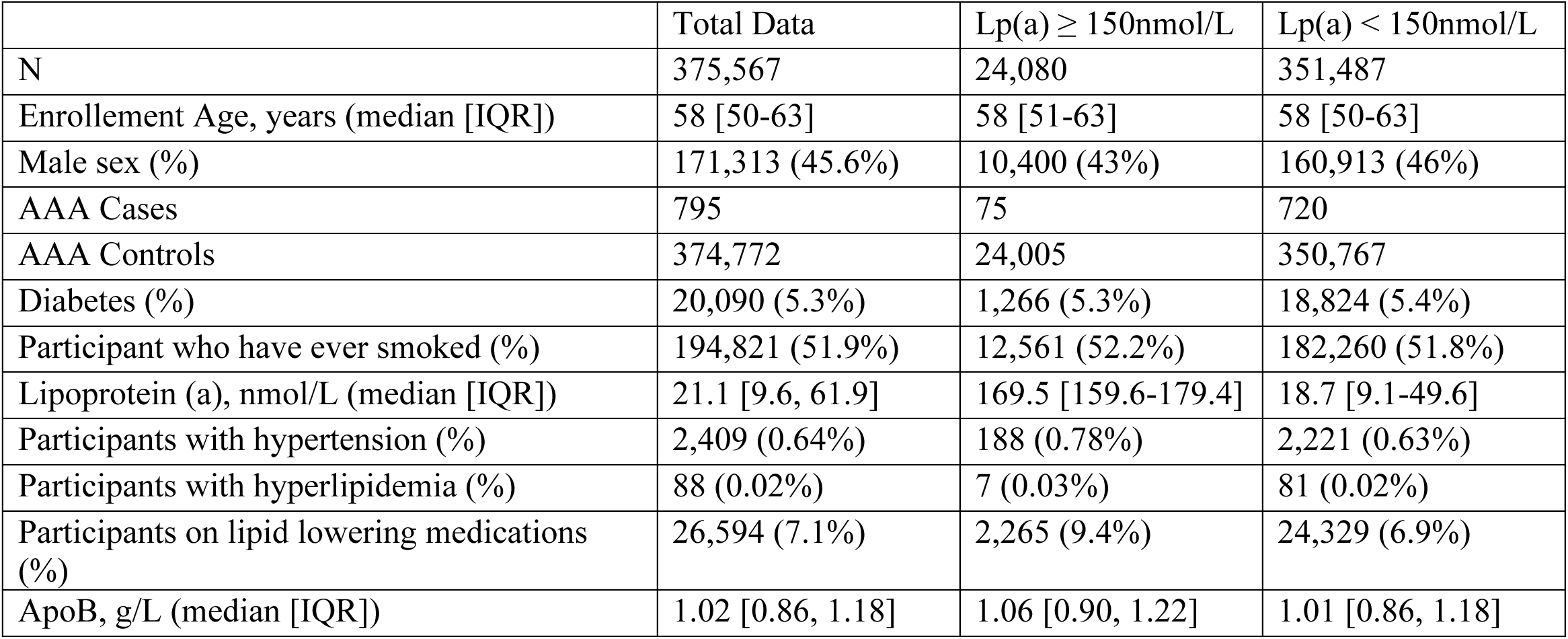
Summary from the UK Biobank Population Analytic Cohort.

| | Total Data | Lp(a) ≥ 150nmol/L | Lp(a) < 150nmol/L |
| --- | --- | --- | --- |
| N | 375,567 | 24,080 | 351,487 |
| Enrollement Age, years (median [IQR]) | 58 [50-63] | 58 [51-63] | 58 [50-63] |
| Male sex (%) | 171,313 (45.6%) | 10,400 (43%) | 160,913 (46%) |
| AAA Cases | 795 | 75 | 720 |
| AAA Controls | 374,772 | 24,005 | 350,767 |
| Diabetes (%) | 20,090 (5.3%) | 1,266 (5.3%) | 18,824 (5.4%) |
| Participant who have ever smoked (%) | 194,821 (51.9%) | 12,561 (52.2%) | 182,260 (51.8%) |
| Lipoprotein (a), nmol/L (median [IQR]) | 21.1 [9.6, 61.9] | 169.5 [159.6-179.4] | 18.7 [9.1-49.6] |
| Participants with hypertension (%) | 2,409 (0.64%) | 188 (0.78%) | 2,221 (0.63%) |
| Participants with hyperlipidemia (%) | 88 (0.02%) | 7 (0.03%) | 81 (0.02%) |
| Participants on lipid lowering medications (%) | 26,594 (7.1%) | 2,265 (9.4%) | 24,329 (6.9%) |
| ApoB, g/L (median [IQR]) | 1.02 [0.86, 1.18] | 1.06 [0.90, 1.22] | 1.01 [0.86, 1.18] |

Based on the thresholds used for the Phase III clinical trials for pelacarsen and olpasiran, clinically elevated levels of Lp(a) are defined as >150 nmol/L.^15,29^ To model this scenario, we dichotomized Lp(a) levels as high (>150 nmol/L) or normal (≤150 nmol/L) and tested the association of high Lp(a) with prevalent AAA. Using this threshold, we found that individuals with clinically elevated levels of Lp(a) (>150 nmol/L) had a 47% increase in odds of having a AAA (OR 1.47; 95% CI 1.15-1.88; P < 0.01) compared to individuals below this threshold.

Finally, to visualize the relationship of Lp(a) to AAA, we plotted the odds ratio in 10nmol/L bins, using the first decile concentration as reference (5.70 nmol/L). Using this model, the effect of Lp(a) on AAA appeared to be linear, as depicted in **Figure 1**.

**Figure 1.**
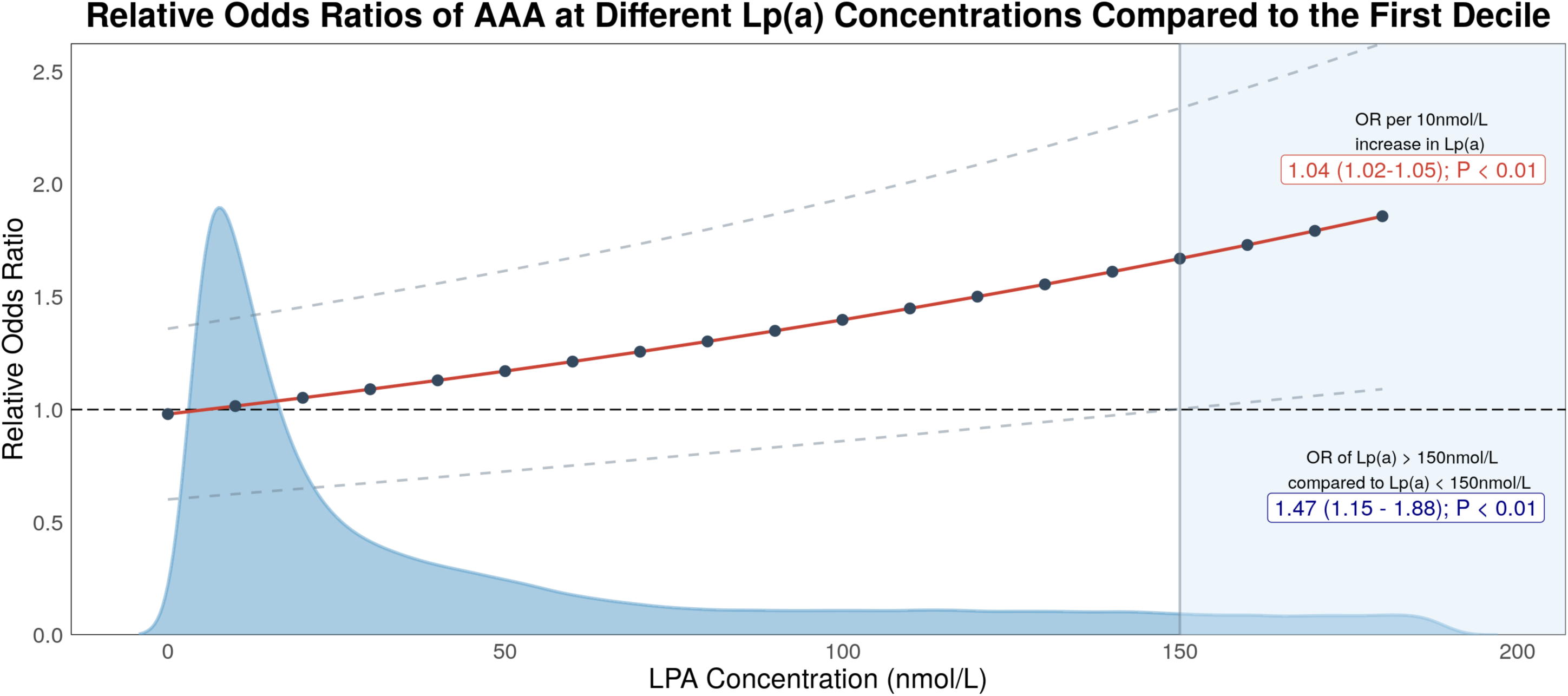
Generalized linear regression model examining the relationship of Lp(a) to AAA compared to the first decile of Lp(a) concentration. Dose-response curve with the relative odds ratio of AAA, scaled by 10 nmol/L compared to the first decile of Lp(a) concentration (5.70 nmol/L). The blue curve represents the population distribution along Lp(a) concentrations. At clinically high levels of Lp(a), defined as greater than or equal to 150 nmol/L, compared to levels below 150 nmol/L, there is a 1.47-fold increase in the odds of AAA (95% CI: 1.15–1.88), as indicated by the blue label. There is also a 4% increase in risk per 10nmol/L increase in Lp(a) (OR 1.04; 95% CI 1.02-1.05; P < 0.01), as highlighted by the red label. The horizontal dotted line represents an OR of 1 at the first decile concentration, with the light blue area of the graph at Lp(a) >= 150 nmol/L representing the relative odds ratio of clinically high levels of Lp(a) compared to the first decile. The red line represents the curve connecting the points at 10 nmol increments, with the grey dashed lines representing the confidence intervals.

### Genetic Causal Inference with Mendelian Randomization

Two-sample MR was used to test the associations of genetically determined Lp(a) and ApoB levels with genetic liability to AAA. Our genetic instruments for Lp(a) (F (mean): 948; r^2^: 0.06) and ApoB (F (mean): 167; r^2^: 0.08) were strongly associated with each respective exposure. After accounting for multiple testing (p < 0.05 ÷ 2 = p < 0.025), both genetically determined Lp(a) and ApoB each showed significant positive associations with genetic liability to AAA in univariate analyses. The odds ratios were 1.20 (95% CI 1.08-1.33, p<0.001) per SD increase in genetically determined levels of Lp(a) (**Figure 2**).

**Figure 2.**
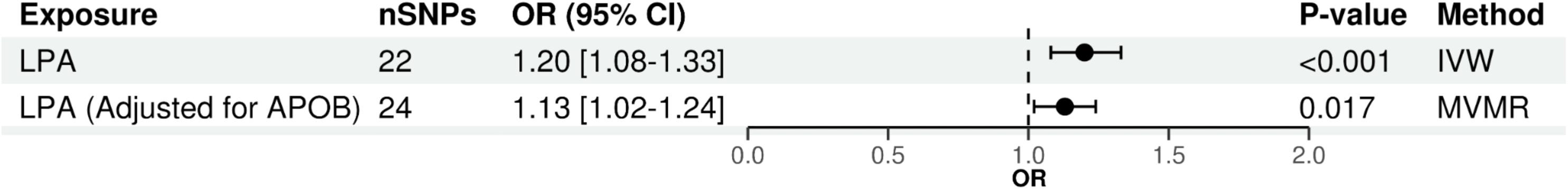
Mendelian randomization analysis exploring the effect of Lp(a) on AAA risk through univariable and multivariable MR methods. This figure highlights the MR analysis results with OR [95% CI] denoting the odds ratio and confidence interval representing 1 SD change in the level of the biomarker. The plot shows the univariable MR analysis using the inverse variance weighted (IVW) method and the multivariable MR (MVMR) analysis, emphasizing a potential risk relationship between Lp(a) and AAA, independent of APOB.

Given that Lp(a) is an ApoB-containing particle, we used MVMR to account for the potential mediating effects of ApoB and estimate the direct effect of genetically determined Lp(a) on genetic liability to AAA, independent of genetically determined ApoB. MVMR confirmed a significant association between increased genetically determined Lp(a) and an increased genetic liability to AAA independent of genetically determined ApoB (OR 1.13 per SD increase in Lp(a); 95% CI 1.02-1.24; P < 0.02) (**Figure 2**).

## DISSCUSSION

We conducted both observational and genetic analyses to investigate the hypothesis that circulating Lp(a) is a risk factor for AAA. We found observational evidence that elevated Lp(a) concentration was associated with a significantly increased risk of AAA, and this effect was independent of traditional cardiovascular risk factors, including ApoB. Genetic analyses further support this association, suggesting a significant association between genetically determined elevated Lp(a) and increased genetic liability to AAA, implying a causal association. These findings have important implications for Lp(a) as both a risk factor and a potential novel therapeutic target for AAA.

Our observational analysis demonstrates a modest increase in risk of AAA per 10nmol/L increase in Lp(a) (OR 1.04; 95% CI 1.02-1.05). However, individuals with clinically elevated Lp(a) concentrations (>150nmol/L) have a substantially higher risk (OR 1.47; 95% CI 1.15-1.88). This heightened risk for AAA parallels that of CAD, where a hazard ratio of 1.63 (95% CI 1.56–1.70) has been reported in subgroups with high Lp(a) without existing ASCVD.^30^ These findings also align with the risk associated with major cardiovascular events (MACE), as genetic studies show a hazard ratio of 1.47 (95% CI 1.43-1.52) for participants with median Lp(a) levels of 146.3 nmol/L compared to those with median levels of 13.6 nmol/L.^31^ These results suggest that the risk conferred by clinically high levels of Lp(a) on AAA mimics the effects seen across other vascular beds, including CAD and MACE, as supported by both observational and genetic studies.

Interestingly, when we visualize our model in increasing 10nmol/L groups of Lp(a), it demonstrates a linear association between higher Lp(a) levels and increased risk of AAA. These findings suggest that the risk of AAA may be graded or proportional to Lp(a) levels, challenging the physiological relevance of current clinical trial thresholds used for outcome enrichment. This largely linear effect, with no evidence of a threshold effect across percentiles, has been previously demonstrated in studies exploring Lp(a) on incidence and risk of ASCVD. As shown by Patel et al., the adjusted hazard ratio of Lp(a) concentrations in the top quintile versus the middle quintile was 1.87 (95% CI, 1.68–2.08), with the same comparison done with LDL showing an adjusted hazard ratio of 2.20 (95% CI, 2.00–2.41).^30^ These findings align with more recent studies, in contrast to previous literature that emphasized the risk associated with Lp(a) is primarily restricted to those with very high concentrations.^32^

The concept of lowering Lp(a) regardless of the baseline is similar to the rationale for lowering LDL cholesterol. Lowering LDL-C significantly reduces cardiovascular events, as evidenced by multiple studies.^33,34^ For instance, early studies from the Cholesterol Treatment Trialists Collaboration found a 22% reduction in CV events with a 1 mmol/L (39 mg/dL) decrease in LDL-C, with benefits seen across all baseline levels, not just high levels of LDL-C.^34^ This evidence has been supported by multiple trials such as PROVE IT, TNT, IMPROVE IT, and SPARCL, all showing that more intensive lowering of LDL-C significantly reduced cardiovascular events irrespective of thresholds.^35–38^ These findings later translated into clinical guidelines, with the 2018 ACC/AHA and 2019 European Society of Cardiology guidelines, emphasizing that ‘lower is better’ for LDL-C.^39,40^ Similarly, Burgess et al. explored the risk reduction of Lp(a) using genetic techniques for CAD, indicating that the clinical benefit of lowering Lp(a) is proportional to the absolute reduction in its concentration. Based on our observation that the Lp(a) effects also appear linear, these findings suggest that substantial reductions in Lp(a) levels, irrespective of baseline levels of Lp(a), may translate into clinically meaningful improvements in AAA outcomes.

Lastly, we observe that the effects of Lp(a) on AAA are largely independent of ApoB. These findings suggest a potential role for Lp(a)-targeting medications in managing AAA in combination with ApoB-lowering medications like statins, which are recommended to reduce the concomitant risk of MACE among individuals with prevalent AAA.^41^ This finding is consistent with prior observations that Lp(a) is markedly more atherogenic than LDL.^42^ Mendelian randomization-derived OR analyses show that for CAD, a 50 nmol/L higher Lp(a)-apoB was 1.28 (95% CI: 1.24-1.33) compared with 1.04 (95% CI: 1.03-1.05) for the same increment in LDL-apoB. Further, the synergistic and independent roles of Lp(a) and ApoB have been highlighted in studies exploring cases of Lp(a) and familial hypercholesterolemia (FH), showing that Lp(a) and FH played a synergistic role in predicting the early onset and severity of CAD.^43^ Additionally, research leveraging data from both UKB and 23andMe has demonstrated that Lp(a) retains a causal effect on MACE even after accounting for LDL-C or apoB.^44^ Here, we demonstrate that this independent relationship of Lp(a) and ApoB extends to other vascular beds and remains true when considering the risk for AAA.

The therapeutic potential of Lp(a)-lowering, particularly in the context of other traditional risk factors for AAA, has remained uncertain.^45^ Ongoing trials such as OCEAN(a) and Lp(a)HORIZON have shown promising results regarding Lp(a) lowering, with a primary focus on cardiac events.^46,47^ Given the absence of current medical management options for AAA, the linear association with no evident threshold effect, and the comparative risk profile to CAD at higher concentrations, Lp(a)-targeted therapy may be a viable option. This study highlights robust observational evidence supporting Lp(a)’s role in AAA pathobiology, reinforced by genetic analyses hinting at its potential causal role, independent of ApoB. These findings suggest that the inclusion of clinical endpoints related to AAA may be valuable to consider in ongoing Lp(a) lowering trials.

## LIMITATIONS

This study should be analyzed within the context of its limitations. First, our observational study relied on data from the UK Biobank, which may not be representative of the broader population due to selection bias.^48^ Additionally, given the relatively low rate of AAA among UK Biobank participants, we utilized prevalent, rather than incident, event analysis, which may have introduced some component of selection bias. However, because Lp(a) levels are not thought to vary substantially over time, this limitation may be less relevant.^49^ Second, there are a number of limitations of Mendelian randomization that stem from its assumptions. While we established that genetic variants selected as instrumental variables were strongly associated with the exposure of interest, MR additionally assumes that genetic variants influence the outcome solely through the exposure and not through other pathways, which may not always hold true in complex biological systems.^26,27^ By selecting genetic variants strongly associated with Lp(a) and ApoB, we satisfied the first assumption. While efforts were made to minimize pleiotropy in genetic analyses, the presence of pleiotropic effects could still influence the results. Sensitivity analyses were conducted to address potential pleiotropy, as presented in **Supplementary Table 3**, but residual confounding may still be present. Third, MR proxies the lifelong effects on normal variation within the physiologic range, in comparison to drug trials which consider potentially more potent effects over a shorter duration which may not represented through this study design.

## CONCLUSION

This study highlights, through both observational and genetic analyses, that Lp(a) confers a risk to AAA that is independent of other cardiovascular risk factors, including ApoB. Given the lack of medical therapy available in the management of AAA, this study underscores the potential role for Lp(a)-targeted therapy in future prevention and treatment of AAA.

## Data Availability

Summary statistics for Lp(a) and ApoB are publicly available at https://pan.ukbb.broadinstitute.org/downloads.
GWAS summary statistics for AAA are publicly available at https://csg.sph.umich.edu/willer/public/AAAgen2023/.

## Nonstandard Abbreviations and Acronyms

AAA: Abdominal Aortic Aneurysms
Lp(a)/LPA: Lipoprotein(a)
ApoB/APOB: Apolipoprotein B
ASCVD: Atherosclerotic Cardiovascular Disease
GWAS: Genome-wide association study
MR: Mendelian Randomization
OR: Odds Ratio
MVMR: Multivariable Mendelian Randomization
UKB: UK Biobank

## ACKNOWLEGEMENTS

This work has been conducted using data from the UK Biobank, a major biomedical database, under application number 70653. The authors sincerely thank the study participants, as well as the staff and researchers of the UK Biobank, for their invaluable contributions and for making the data available for this study.

## SOURCES OF FUNDING

This work was supported by the NIH NHLBI R01HL166991. MGL was supported by the Doris Duke Foundation (Award 2023-0224) and US Department of Veterans Affairs Biomedical Research and Development Award IK2-BX006551.

## DISCLOSURES

SMD receives research support to University of Pennsylvania from RenalytixAI and in kind support from Novo Nordisk, both outside the scope of this research. MGL receives research support to University of Pennsylvania from MyOme, outside the scope of this research. The content of this manuscript does not represent the views of the Department of Veterans Affairs or the United States Government.

## Supplemental Material

Tables S1–S5

